## Supplementary Information for "A Digital Biomarker for Identifying Changes in Daily Activity Patterns"

In this section, we provide the supplementary evidence necessary to support our claims.

*A. Study Cohort Demographics*

Here we detail the distribution of diagnoses within the study cohort, as well as the variation in year of birth within the study cohort.

TABLE I: Diagnoses for the study cohort

| Dementia Type | %Male | %Female | %Total |
|---|---|---|---|
| **Alzheimer's Disease** | 34% | 36% | 70% |
| **Dementia with Lewy Bodies** | 3% | 0% | 3% |
| **Frontotemporal Dementia** | 1.5% | 1.5% | 3% |
| **Parkinson's Disease Dementia** | 7% | 0% | 7% |
| **Vascular Dementia** | 5% | 0% | 5% |
| **Other/Not specified** | 5% | 7% | 12% |

TABLE II: Year of birth statistics for the study cohort

| Total Count | Missing | Mean | Std Dev | Min | Max |
|---|---|---|---|---|---|
| **72** | 1(1.5%) | 1941 | 8.24 | 1927 | 1962 |

*B. Minder Study Inclusion and Exclusion Criteria*

Here we detail the comprehensive criteria by which an individual may be included or excluded from the Minder study.

*1) Inclusion Criteria:* People living with dementia must meet the following criteria to be included in the Minder study:

- Have an established dementia of diagnosis or mild cognitive impairment by specialist assessment.
- Be male or female over the age of 50 years old.
- Have sufficient functional English to allow completion of the assessment instruments.
- If lacking in capacity, participants must have a personal consultee representative.
- Have a study partner over the age of 18, with sufficient functional English, willing and able to provide informed consent and who has known the person living with dementia for at least 6 months and is able to attend research assessments with them.

*2) Exclusion criteria:* People living with dementia who meet the following criteria are excluded from the Minder study:

- Are in receipt of any investigational drug within a 30-day period prior to consenting.
- Have an unstable mental state including severe depression, severe psychosis, agitation, and anxiety whose medication was changed over the last 4 weeks.
- Have a severe sensory impairment.
- Have active suicidal ideation.
- Require regular elective hospital admission for monitoring of their physical health.
- Are receiving treatment for a terminal illness.
- Have study partners who are unable to communicate verbally or provide written informed consent.
